# Investigating the Effectiveness of Breast Cancer Supplemental Screening Considering Radiologists’ Bias

**DOI:** 10.1101/2020.12.16.20248373

**Authors:** Sevda Molani, Mahboubeh Madadi, Donna L. Williams

## Abstract

Breast density is known to increase breast cancer risk and decrease mammography screening sensitivity. Breast density notification laws (enacted in 38 states as of September 2020), require physicians to inform women with high breast density of these potential risks. The laws usually require healthcare providers to notify patients of the possibility of using more sensitive supplemental screening tests (e.g., ultrasound). Since the enactment of the laws, there have been controversial debates over i) their implementations due to the potential radiologists bias in breast density classification of mammogram images and ii) the necessity of supplemental screenings for all patients with high breast density. In this study, we formulate a finite-horizon, discrete-time partially observable Markov chain (POMC) to investigate the effectiveness of supplemental screening and the impact of radiologists’ bias on patients’ outcomes. We consider the conditional probability of eventually detecting breast cancer in early states given that the patient develops breast cancer in her lifetime as the primary and the expected number of supplemental tests as the secondary patient’s outcome. Our results indicate that referring patients to a supplemental test solely based on their breast density may not necessarily improve their health outcomes and other risk factors need to be considered when making such referrals. Additionally, average-skilled radiologists’ performances are shown to be comparable with the performance of a perfect radiologist (i.e., 100% accuracy in breast density classification). However, a significant bias in breast density classification (i.e., consistent upgrading or downgrading of breast density classes) can negatively impact a patient’s health outcomes.

## 1 Introduction

Breast cancer is a leading cause of premature mortality among women in the U.S. [39]. According to the American Cancer Society (ACS), the risk of death from breast cancer is approximately 1 in 38 [5]. The ACS estimates that 42,170 women will die from breast cancer in 2020 [5]. Breast cancer screening tests, the most common of which is mammography, can help with detecting breast cancers in early stages and thereby reducing the breast cancer mortality risk by treating patients when they have a higher survival chance. However, due to the imperfect nature of mammography screening, there is always some associated false positives and false negatives risks. False negative rates are especially higher in women with dense breasts due to the reduced sensitivity of mammography caused by the masking effect of high density breast tissue.

Breast density is defined as the prevalence of fibroglandular tissue in the breast. The Breast Imaging Reporting and Data System (BI-RADS) classification system classifies breast tissue density into four categories: almost entirely fatty which includes less than 25 percent glandular tissue (class 1), scattered fibroglandular which includes approximately 25-50 percent glandular tissue (class 2), heterogeneously dense which includes approximately 51-75 percent glandular tissue (class 3), and extremely dense which includes more than 75 percent glandular tissue (class 4) [6].

Breast density is associated with an increased risk of breast cancer [35]. It is well-established that tumors in dense breasts may progress more rapidly than those in fatty breasts [35]. Previous studies have shown that the relative risk of breast cancer associated with breast density is substantially higher than other risk factors such as breast cancer family history and menstrual and reproductive risk factors [24]. The reported odds ratio for developing breast cancer for the most dense compared with the least dense breast tissue categories ranges from 1.46 [18] to 6.0 [22].

Additionally, higher breast density can significantly reduce the mass detection rate since the normal tissues in dense breasts appear as bright areas in mammography. Due to the lower sensitivity of screening mammography in women with dense breasts, the cancer is more likely to remain undetected.

Breast density is a dynamic risk factor and typically decreases as a patient becomes older [27, 48, 51]. Younger women (especially those in pre-menopausal status) are more likely to have dense breasts [1]. According to Mandelblatt et al. [32], 58.8% of women aged 40-49 have highly dense breasts, while this percentage decreases to 42.7% and 31.1% for women aged 50-64 and 65-74, respectively.

Breast density notification laws have been enacted in 38 states in the U.S. (as of August 2020) to mitigate the increased breast cancer risk in women with high breast density which is partially caused by the masking effect of dense breasts in screening mammography. These laws generally require physicians to notify patients with high breast density (classes 3 and 4) of their increased risk of breast cancer compared to women with low breast density. Moreover, in some states, the breast density notification law requires physicians to inform women with high breast density that adjunctive screening tests such as breast ultrasound and magnetic resonance imaging (MRI) may benefit them. Breast ultrasound uses high-frequency sound waves to make an image of breast tissue and as a result, has higher sensitivity than mammography in women with dense breasts. MRI uses intravenous contrast solution injection to produce 3-dimensional images of breast tissue.

Since the emergence of breast density notification laws, there have been controversial debates on the potential unintended consequences of the laws as well as their implementation quality. It is believed that supplemental screening may result in an increased number of unnecessary supplemental screening tests and biopsies as well as patients’ overdiagnosis (i.e., detection of a cancer that would not have become clinically apparent over the patient’s lifetime if left undetected). In addition, inter and intra-variabilities in breast density classification by radiologists raised some concerns since it results in patients’ breast density misclassification (e.g., classification of a patient to a breast density category different from her true BI-RADS breast density class) [20]. According to Bahl et al. [11], the percentage of mammogram images reported as dense decreased after the enactment of breast density notification laws. This reduction happens as radiologists may downgrade their assessment of density to avoid reporting requirements. On the other hand, there have been controversies that radiologists may upgrade their assessments so that supplemental screening can be ordered and their liability is minimized [11].

Currently, there is not a consensus among different health agencies in the U.S. regarding the necessity of supplemental screening in early breast cancer detection for women with high breast density. The American College of Radiology (ACR) advocates the use of ultrasound as an adjunctive screening test in women with high breast density [25]. However, according to the ACS report, there is not enough evidence to make a recommendation for or against supplemental screening in women with dense breasts [3]. The U.S. Preventive Services Task Force (USPSTF) and the American College of Physicians (ACP) state that the current evidence is not sufficient to support the recommendation of supplemental screening [47, 50].

### 1.1 Relevant literature

Markov models have been previously used to evaluate/optimize breast cancer screening and treatment strategies. Nohdurft et al. [38] formulated a Markov decision process (MDP) to derive optimal surgery decisions for women with breast cancer. Chhatwal et al. [15] developed a finite-horizon discrete-time MDP to provide patient-specific recommendations for breast biopsy based on the patient’s mammographic features. Alagoz et al. [2] formulated a finite-horizon discrete-time MDP to optimize the post-mammography diagnostic decisions (choosing between biopsy or short-interval follow up mammogram) based on mammogram test findings. Ayvaci et al. [10] developed an MDP model to optimize the risk-sensitive diagnostic decisions after a mammography exam. In their study, the radiologist can select from biopsy, short-term follow-up, and routine mammography while considering a patient’s preference to maximize the quality-adjusted survival duration. In another study, Ayvaci et al. [9] investigated the impact of budgetary restrictions on breast biopsy decisions by developing a constrained MDP. Çağlayan et al. [13] developed a Markov framework to model breast cancer progression incorporating different risk factors such as gene mutations and family history of breast and ovarian cancer. They then identified the optimal and most cost-effective population screening strategies.

As mammography is not perfect and may not reflect the true health status of a patient, some studies used partially observable Markov models in assessing/optimizing breast cancer screening policies. Maillart et al. [31] formulated a partially observable Markov chain (POMC) model to compare different breast cancer screening policies in terms of lifetime breast cancer mortality risk and the total expected number of mammograms. Ayer et al. [7] formulated a partially observable Markov decision process (POMDP) to determine optimal personalized mammography screening policies maximizing a patient’s quality adjusted life years (QALYs). In another study, Ayer et al. [8] developed a POMDP framework to analyze the importance of heterogeneity in women’s adherence to mammography screening policies. Madadi et al. [30] developed a discrete-time POMC model to evaluate mammography screening policies in terms of the expected QALYs and lifetime breast cancer mortality risk while incorporating the uncertainty in women’s adherence behaviors. Molani et al. [34] developed POMC models to quantify the age and stage-specific overdiagnosis risks while considering the uncertainty in a patient’s adherence behavior. Cevik et al. [14] proposed a POMDP model to maximize the total expected QALYs of a patient when there is a constraint on the number of mammograms the patient can undergo. Sandikci et al. [44] formulated a POMDP model to determine the optimal breast cancer screening policies considering patients’ breast density. Otten et al. [41] formulated a finite horizon discrete-time POMDP to optimize and personalize breast cancer follow-up.

In this paper, we develop a POMC model to investigate the impact of radiologists’ bias on patients’ health outcomes under the breast density notification law. The patients’ health outcomes include the conditional probability of detecting breast cancer in early and advanced cancer states given the patient develops breast cancer in her lifetime and the total expected number of supplemental screening tests a patient undergoes in her lifetime. We consider the conditional probability of detecting breast cancer in early states as the primary health outcomes since detecting breast cancer in early states, where the patient has higher survival chances, is the main purpose of cancer screening programs [37].

To the best of our knowledge, in the operations research literature, Sandikci et al. [44] work is the only study that explicitly models breast density as a significant breast cancer risk factor. Our study, however, is different from Sandikci et al.’s work in several aspects: 1) We consider the conditional probability of detecting breast cancer in early and advanced states given that the patient develops cancer in her lifetime as the main patient’s health outcome. To the best of our knowledge, this is the first study in the literature to consider these patients’ health outcomes. 2) We investigate the impact of radiologists’ bias in density classification of mammogram images on patients’ health outcomes. This is done by modeling breast density as a partially observable variable. In Sandikci et al.’s work, however, breast density is assumed to be fully observable (i.e., radiologist’s evaluation of breast density perfectly correlates with the patient’s actual density). 3) In this study, we use sequential mammography screening data of 436 patients from Louisiana Cancer Prevention and Control Programs [28] to estimate the dynamics of breast density to more accurately model the breast cancer risk dynamics caused by potential changes in breast density.

The remainder of this paper is as follows. In Section 2, we formulate a POMC model to calculate the patient’s health outcomes. Section 3 presents parameter estimations and model validation. Numerical results and sensitivity analyses are presented in Section 4. Finally, we summarize and conclude in Section 5.

## 2 Model formulation

A discrete-time finite-horizon partially observable Markov chain (POMC) is developed to model breast cancer natural history and breast density dynamics. A POMC is used as the imperfect nature of mammography tests (i.e., possibility of receiving false positives and false negatives) as well as the possibility of breast density misclassification by radiologists prevent the patient’s true state to be fully observable to the decision maker.

The primary patients outcome measure is the conditional probability of detecting cancer in early/advanced states given that the patient develops breast cancer in her lifetime. More specifically, we focus on the population of patients who would develop breast cancer at some point in their lifetimes and their cancer eventually becomes symptomatic if not detected through screening tests. The latter assumption is made to rule out the over-diagnosed cases as for these cases, the detection of cancer is not favorable. Note that early detection of cancers which will eventually become problematic is the main incentive of screening programs [37].

Obviously, the more aggressive a screening strategy is (more frequent and multiple screening modalities), the higher is the chance of detecting cancer in early states where it is more likely to be treated. However, there are disutilities associated with screening tests that adversely impact a patient’s quality of life. Therefore, there is a trade-off between detecting cancer in early states and the discomfort of undergoing aggressive frequent screenings. As such, we consider the total expected number of supplemental screening tests a patient would undergo in her lifetime as another patients’ outcome to investigate the trade-ff.

We estimate the conditional probability of eventually detecting breast cancer in early and advanced state in Section 2.1 and the total expected number of supplemental screening tests a patient undergoes in her lifetime in Section 2.2. The following is the list of notation used in the proposed model. Note that vectors and matrices notations are in bold.

*t* Time periods, *t* = 0, 1, 2 …, *T*.

*s* Patient’s core state; Specifically, *s* = (*h, d*) *∈* Ω = *H* × *D* represents the patient’s underlying state where *h ∈ H* and *d ∈ D* denote the patient’s core health and breast density states, respectively. The health state set *H* includes three partially observable states of cancer-free (state 0), early breast cancer (state 1), and advanced breast cancer (state 2) and one fully observable state of death due to breast cancer or other causes (state 3). Specifically, we refer to the partially and fully observable health state sets as *H*_1_, and *H*_2_, respectively, i.e., *H* = *H*_1_ *∪ H*_2_. We denote the subsets of patient’s core states for which *h ∈ H*_1_ and *h ∈ H*_2_ by Ω_1_ and Ω_2_, respectively. Moreover, set *D* includes four BI-RADS density classes as discussed in Section 1, i.e., *D* = {1, 2, 3, 4}.

***β***_*t*_ A vector of length |Ω_1_| representing the patient’s belief state at the beginning of period *t*. Specifically, ***β***_*t*_(*s*) denotes the probability that the patient is in partially observable state *s* = (*h, d*), *h ∈ H*_1_ at the beginning of period *t*.

***P***_*t*_ Underlying transition probability matrix capturing the natural history of breast cancer and breast density dynamics. That is, ***P***_*t*_(*s*^*′*^|*s*) represents the probability that a patient will be in state *s*^*′*^ = (*h*^*′*^, *d*^*′*^) at time *t* + 1, given that she is in state *s* = (*h, d*) at time *t*.

*a*_*t*_ Prescribed action at time *t*, where possible actions include *wait* and *mammography*, denoted by *W* and *M*, respectively. A patient classified in a high density class undergoes a supplemental screening following a negative mammogram, for which case the action is denoted by *B*. Let *A* denote set of all possible actions; we have *a*_*t*_ *∈ A* = {*W, M, B*}.

*o*_*t*_ Observation received at time *t* which includes both breast density classification by the radiologist and screening result. Specifically, *o*_*t*_ = (*θ, δ*), where *θ ∈* Θ = *D* and *δ ∈* Δ^*a*^ respectively denote the assigned breast density class and screening result. For notation brevity, we define density observation subsets <u>Θ</u> = {1,2} and 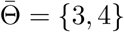. Clearly, we have 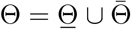. Observations are received only if the prescribed action is to undergo a mammogram. Specifically, when patient undergoes only a *mammography*, the possible test results are negative and positive mammogram, respectively denoted by *M* ^*−*^ and *M* ^+^. That is, Δ^*M*^ = {*M* ^*−*^, *M* ^+^}. When a mammogram is *accompanied with a supplemental* test (i.e., *a*_*t*_ = *B*), possible observations are *M* ^*−*^&*S*^*−*^ and *M* ^*−*^&*S*^+^ which respectively represent a negative mammogram followed by a negative and a positive supplemental test, i.e., Δ^*B*^ = {*M* ^*−*^&*S*^*−*^, *M* ^*−*^&*S*^+^}. If the action is *wait*, no observations will be received.

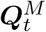 Breast density information matrix, where 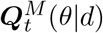 denotes the probability of a patient with true density *d* be classified in density class *θ* upon action *a*_*t*_ = *M*.

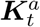 Health information matrix, where 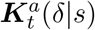 represents the probability of observation *δ ∈* Δ^*a*^ when action *a*_*t*_ *∈* {*M, B*} is taken and the patient’s true state is *s* = (*h, d*) at time *t*. Note that the health observation probability matrix for the case that the action is a mammogram is a function of breast density due to the masking effect of high breast density on mammogram sensitivity.

***η***_*t*_ A vector of length |Ω_1_| representing the probabilities of a patient showing symptoms in period *t*. Specifically, ***η***_*t*_(*s*) is the probability of showing symptoms in state *s* at time *t*.

The one-period sample path of the breast cancer detection process under the notification law is presented in Figure 1. At each period, depending on the prescribed action and possible subsequent observations, patient takes a different path. We assume that after receiving a positive screening result (either a mammogram or a supplemental test), the patient undergoes a biopsy test. Biopsy is assumed to be perfect as its true positives rate is very close to 1 [42]. According to the U.S. Department of Health & Human Services report, biopsy could be considered a test without measurement error [16]. We assume that the probability of both developing cancer and showing symptoms in one period (one year) is zero. That is, cancers can only show symptoms in a period when the patient is in a cancer state at the beginning of that period. Moreover, we assume that breast cancer cannot spontaneously (without treatment) regress [23].

**Figure 1:**
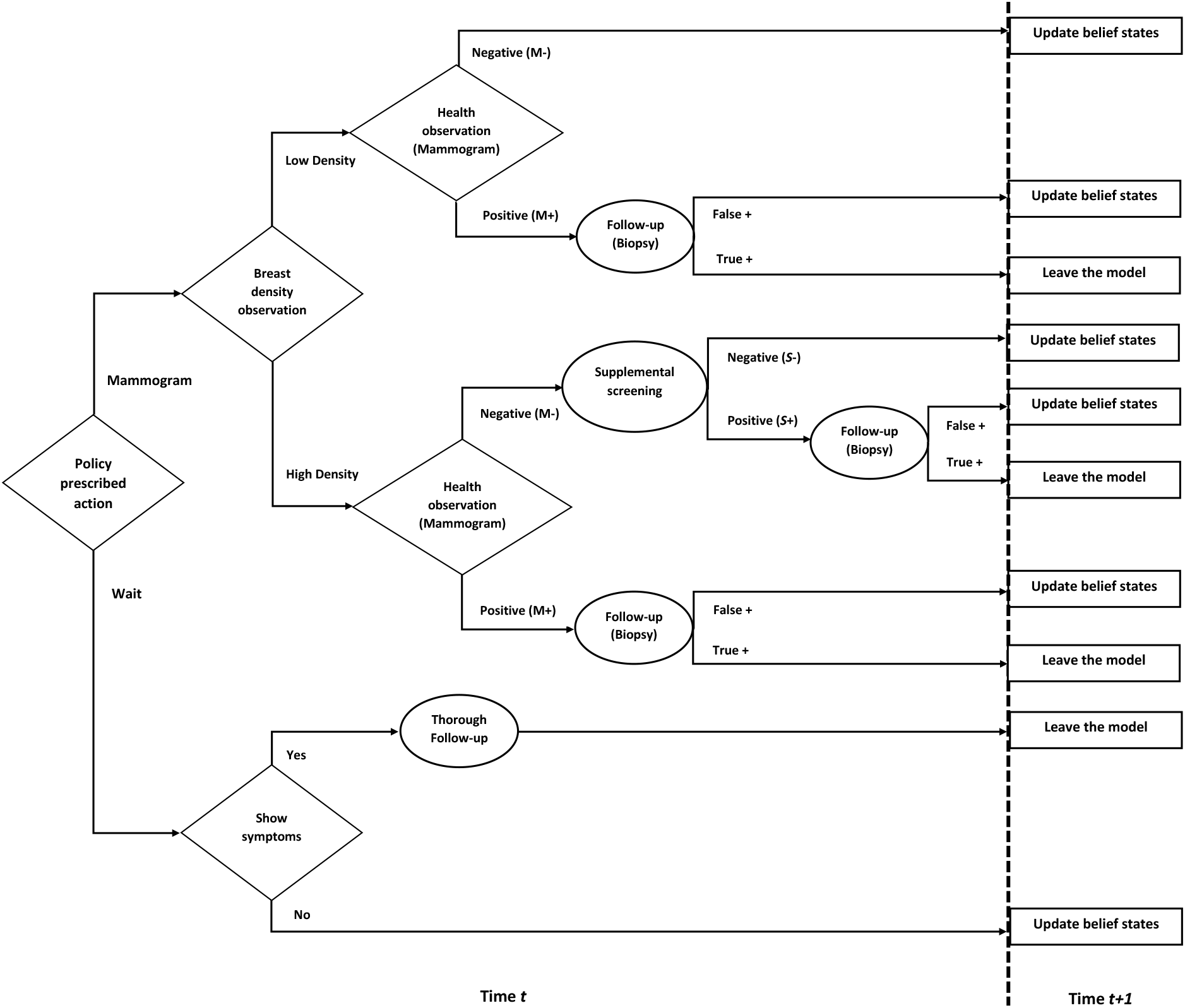
One-period sample paths of the breast cancer detection process when supplemental screening is administered. Note that under action *wait*, symptoms can only happen when the patient is in a cancer state.

At each period, the patient’s belief state is updated based on the action taken and possible observations received. Under action *a*_*t*_, observation *o*_*t*_ and assuming that the patient belief at the beginning of period *t* is ***β***_*t*_, Equation (1) calculates the patient’s updated belief (***ν***) at time *t* + 1:

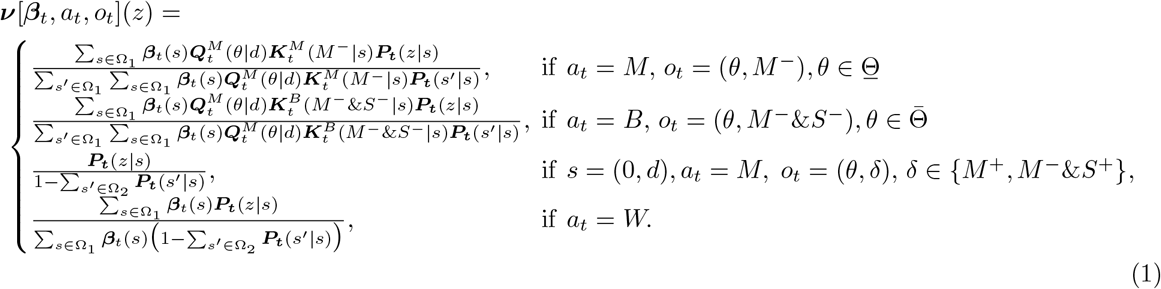

The first and second cases in Equation (1) represents the case when the patient undergoes a prescribed mammogram, receives a negative mammogram result, and is classified into the low and high density class by the radiologist, respectively. In the latter case, the patient undergoes a supplemental screening test and receives a negative result. We use Bayes rule to update the patient’s belief state in these two cases. In the second case where the patient undergoes both mammography and supplemental tests, the joint information from both tests is used to update the patient’s belief state. Note that given the patient health and breast density state, we assume that density observation and test results observations are independent. The third case represents a false positive and consists of two different situations: 1) a false positive mammogram result, and a negative mammogram followed by a false positive supplemental test. Note that in both cases, the true state of the patient (cancer-free) is revealed by a follow-up biopsy. In these cases, the patient’s belief is updated by accounting for possible cancer development from the cancer-free state (*s* = (0, *d*)). The fourth case represents the situation where the action is *wait*. In this case, no observation is received and breast cancer natural history and dynamics of breast density are used to update the patient’s belief state. The term 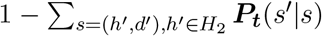 in the third and fourth cases represents the probability that a patient in state *s* survives period *t*.

### 2.1 Probability of detecting cancer in early and advanced states

Let 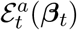 and 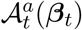 respectively denote the probability of eventually detecting a cancer in early and advanced cancer states when the patient belief state at the beginning of period *t* is ***β***_*t*_ and action *a*_*t*_ is taken. Note that we only consider the cancer population whose cancer will eventually show symptoms. That is, we exclude the over-diagnosed cases whose cancer may never show symptoms or cause any problems. Equations (2) calculates the probability of eventually detecting the cancer in early states when the patient’s belief state is ***β***_*t*_ and the prescribed action in period *t* is *wait*:

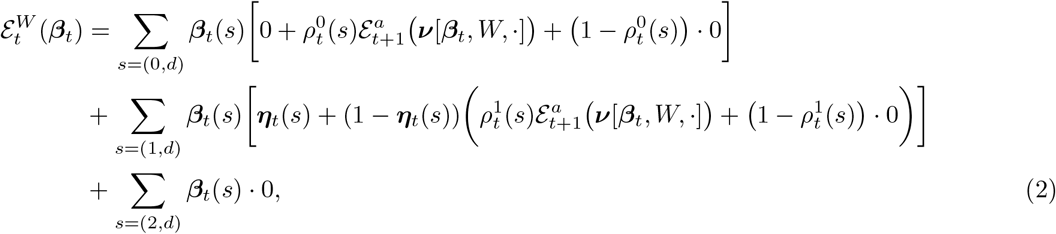

where 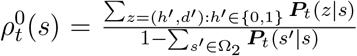 is the probability of remaining in the healthy states or transitioning to early cancer states in period *t* given that the patient is healthy (i.e., *s* = (0, *d*)) at the beginning of period *t* and survives the current period. Additionally, 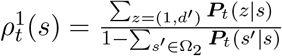 is the probability of remaining in early cancer state (i.e., *s* = (1, *d*)) in period *t* given that the patient survives the current period.

The logic of Equation (2) is as follows: If the patient is in a healthy state at time *t*, the probability of cancer detection in the current period is zero. The future probability of eventually detecting cancer in early state is conditioned on patient’s surviving and not transitioning to advanced states in the current period. That is because of the assumption that cancers cannot spontaneously regress. The probability of such event is 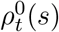 and in such a case, the future probability of cancer detection in early states is 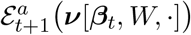. If the patient transitions to an advanced cancer state (with probability 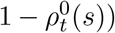), the probability that the cancer will be eventually detected in early states is zero. If the patient is in an early cancer state at the beginning of period *t*, the cancer may show symptoms with probability ***η***_*t*_(*s*). In this case, the follow-up tests will reveal the cancer and the patient leaves the model. If the cancer remains undetected in the current period (which happens with probability 1 *−* ***η***_*t*_(*s*), *s* = (1, *d*)) and the patient remains in early cancer state (which happens with probability 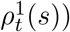), the cancer might be eventually detected in an early state with probability 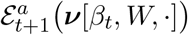. If the patient is in an advanced cancer state at the beginning of period *t*, the cancer can never be detected in an early state since we assume that no cancer regression can occur.

Equations (3) calculates the probability of eventually detecting the cancer in advanced states when the patient’s belief state is ***β***_*t*_ and the prescribed action in period *t* is *wait*:

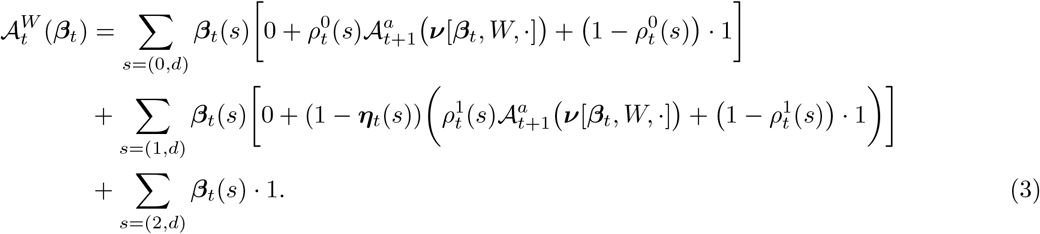

The logic of Equation (3) is as follows: If the patient is in a healthy state at the beginning of period *t*, the immediate probability of detecting cancer in an advanced state is zero. If the patient stays in a healthy or transitions to an early cancer state conditioning that she has survived the current period (which occurs with probability 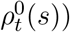), she might eventually be detected in an advanced cancer state with probability 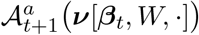. However, if she transitions to an advanced cancer state in the current period, with certainty she will eventually be detected in advanced cancer states. If the patient is in an early cancer state at the beginning of period *t*, her cancer needs to remain undetected in the current period (which happens with probability 1 *−* ***η***_*t*_(*s*)) in order to be later detected in an advanced state. In such a case, if she remains in the early cancer state, the future probability of detecting cancer in an advanced state is 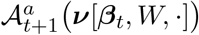, and if she transitions to an advanced state, the corresponding probability is one. Finally, if the patient is in an advanced cancer state, she will eventually be detected in an advanced state with certainty.

Equations (4) and (5) respectively present the probability of eventually detecting the cancer in early and advanced states starting from belief state ***β***_*t*_ at the beginning of period *t* when the prescribed action is a screening *mammogram* with a possible subsequent supplemental test. Note that in compliance with the breast density notification laws, when the prescribed action is a *mammogram*, the patient may take different paths depending on their observed breast density class.

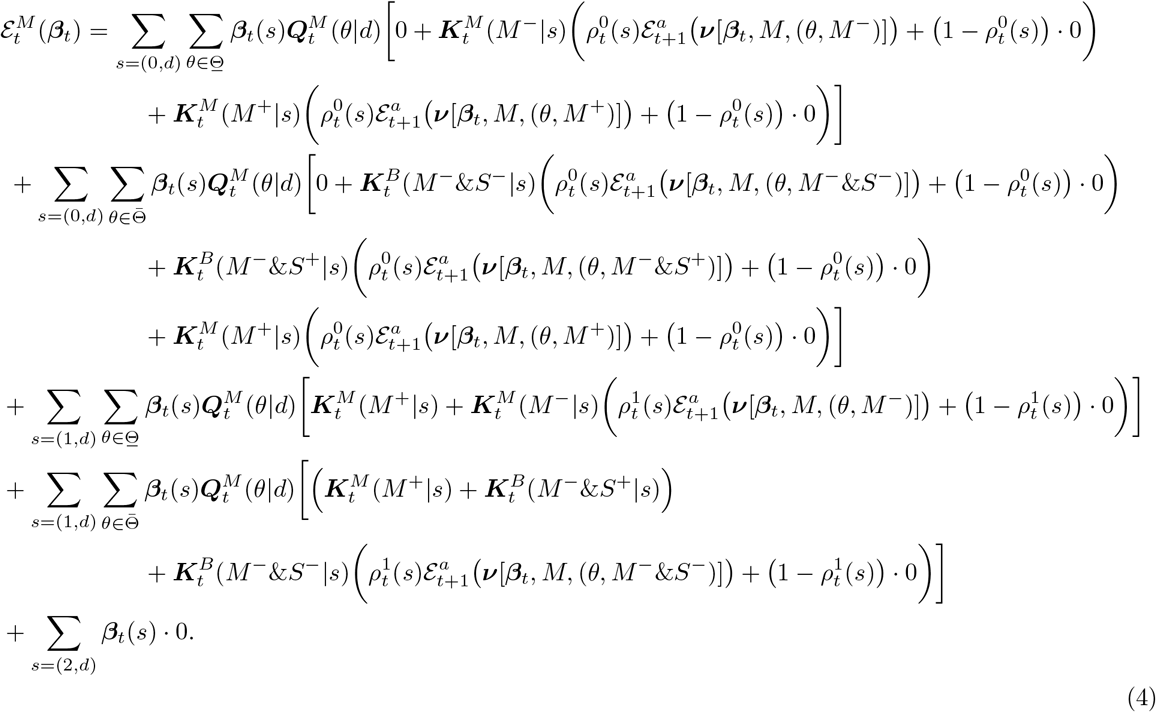

Equation (4) emerges from the following logic. If the patient is healthy, the probability of cancer detection in the current period is zero. If she is classified in the low density class, which occurs with probability 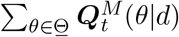, she only receives a mammogram test. The mammogram test result might be a true negative or a false positive with corresponding probabilities of 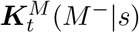 and 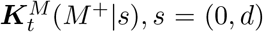. In either case, if the patient remains in the healthy state or proceeds to early cancer state, her belief state is updated and the future probability of cancer being detected in an early state is calculated. Note that we assume in case of a false positive, further examination (i.e, biopsy) reveals that the patient is healthy. If the patient transitions to an advanced cancer state, her future probability of being detected in an early cancer state is zero. When the patient is classified into a high breast density class (with probability 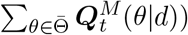, she may undergo a supplemental screening if the mammogram result is negative. Possible outcomes in such a case are a negative mammogram followed by a negative supplemental test (true negative with probability 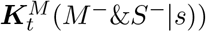, negative mammogram followed by a positive supplemental test (false positive with probability 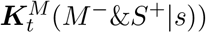, or a positive mammogram (false positive with probability 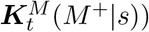. In any of these cases, if the patient does not proceed to advanced cancer states, her belief state is updated based on the received observations and her future probability of being detected in early states is calculated. However, if the patient proceeds to advanced cancer states, the cancer will never be detected in early states.

If the patient is in early cancer states at the beginning of period *t*, her cancer may be detected in the current period through screening tests. Specifically, if the patient is classified as a low and high breast density patient, the cancer may be detected in the current period with probability 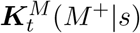 and 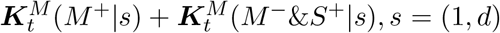, respectively, in which case the patient leaves the model. However, if the screening does not reveal the cancer, which occurs with probabilities 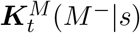 and 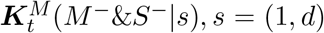 when the patient is classified as low and high density class, respectively, the patient belief is updated based on the sensitivity of screening test(s) that the patient has undergone and the future probability of the cancer being detected in early state is calculated. Finally, if the patient is in advanced cancer states at the beginning of period *t*, the probability that she eventually be detected in an early cancer state is zero.

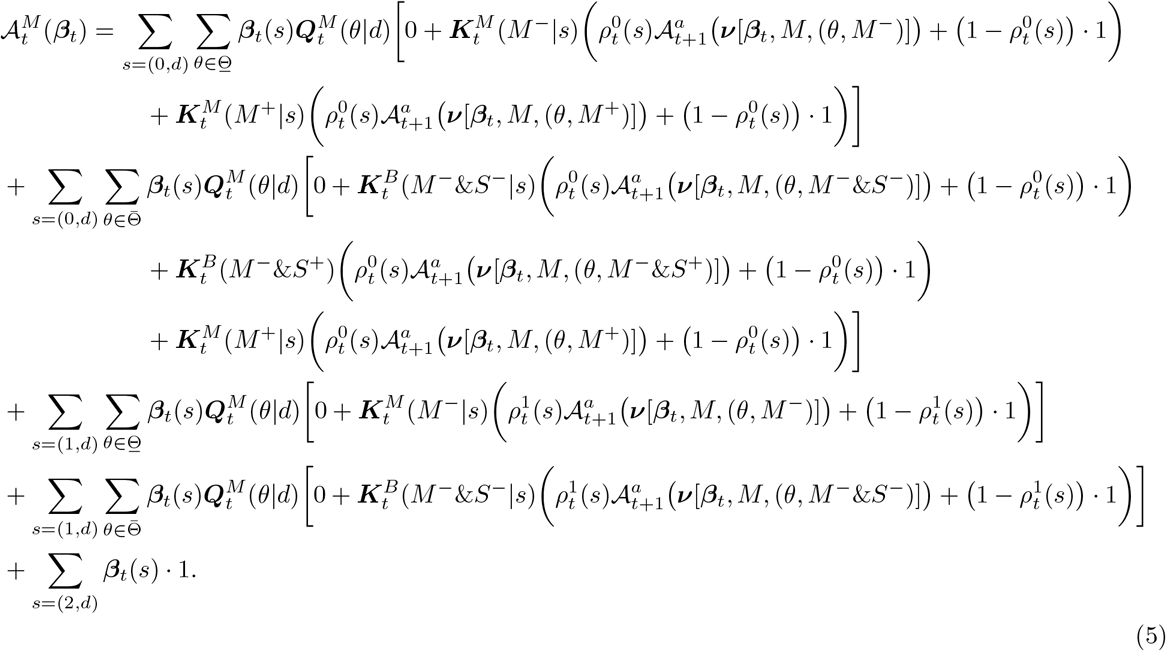

Equation (5) follows a logic similar to that of Equations (3) and (4) and thus is omitted for brevity.

For the boundary conditions, a healthy patient or a patient in early cancer states can be eventually detected in either early or advanced cancer states. The probability of such events are estimated using cancer progression rates and probability of showing symptoms after period *T*. For a patient in advanced cancer states at time *T*, the cancer will eventually be detected in the advanced states. Let *γ*_*E*_(*s*) and *γ*_*A*_(*s*) respectively denote the probability of eventually detecting cancer in early and advanced states for a patient in state *s* at time *T*. We have

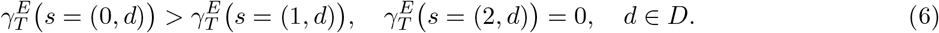

The probabilities 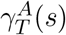 are then calculated using the fact that 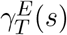 and 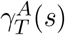 are complementary. Recall that we assume 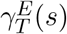 and 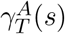 are complementary to exclude over-diagnosed cases.

### 2.2 Expected number of supplemental screenings

Let 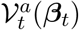 denote the expected number of supplemental screenings a patient undergoes in her remaining life years when at the beginning of epoch *t*, she is in belief state ***β***_*t*_ and the prescribed action is *a*_*t*_. Equations (7) and (8) calculate 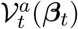 for actions *wait* and *mammogram*, respectively. Note that we assume when a patient shows symptom a thorough follow-up (subsequent supplemental test and biopsy) will be administered. When the prescribed action is wait, the patient may undergo a supplemental screening if the cancer shows symptoms. Specifically, when the patient is in state *s* = (*h, d*) *∈* Ω_1_, she may develop symptoms with probability ***η***_*t*_(*s*). If the follow-up mammogram result is negative, she will undergo subsequent supplemental test. If the follow-up mammogram is positive, the patient does not receive supplemental test and leaves the model. However, if she does not show any symptoms (with probability 1*−****η***_*t*_(*s*)), she proceeds to the next period with probability 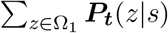 and the future expected number of supplemental screening is 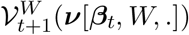.

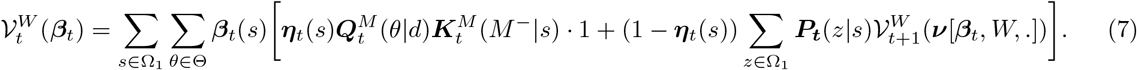

When the prescribed action is a mammogram, the patient receives a supplemental screening only if she is classified as a high-density patient in the mammogram screening and the mammogram result is negative. In such cases, she receives an immediate cost of 1. If the supplemental test result returns positive and the patient is actually in a cancer state, she leaves the model. In any other case, the patient belief state is updated based on the screening test(s) and the corresponding observation she has received and her future cost-to-go is calculated.

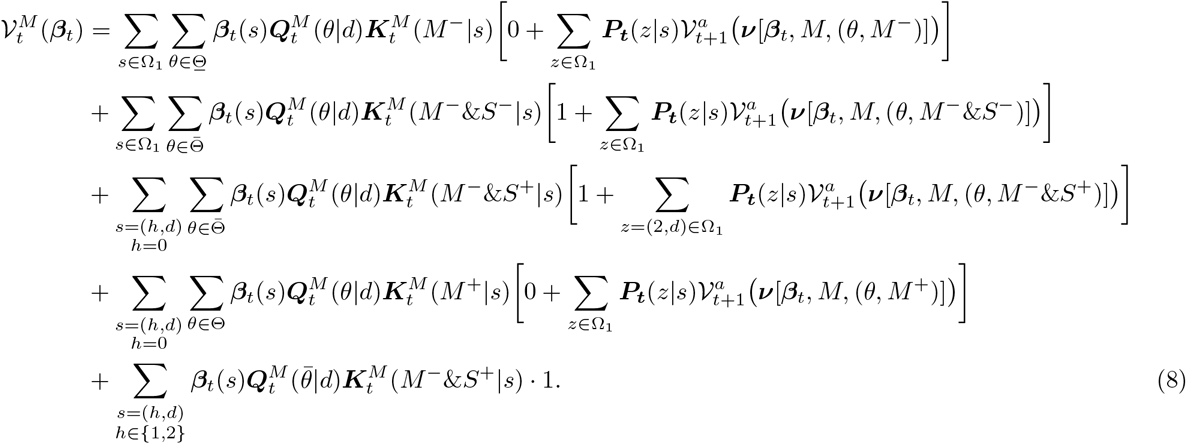

To calculate the probability of detecting cancer in early and advanced states and the expected number of supplemental screening, we first enumerate all possible sample paths a patient may undergo for a particular screening policy. We then calculate 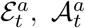 and *𝒱*_*t*_ for all occupancy distributions on these sample paths using Equations (2) through (8), starting from *t* = *T* and moving backward to *t* = 0 from each possible ending distribution.

## 3 Parameters estimation and model validation

The data sources used to estimate the parameters of the proposed model are presented in Table 1. Following the recommended screening policies in the U.S., we use age 45 and 75 (corresponding to *t* = 0 and *t* = *T* = 30) as the earliest and latest ages that a patient undergoes a breast cancer screening test. The start age of 45 is considered based on the new ACS policy recommendation and the fact that the risk of developing breast cancer is very small in women younger than 45 [3]. Moreover, we assume no screening is administered after age 75 since the risks associated with breast cancer screening outweigh its benefits in women older than 75 [29].

**Table 1:**
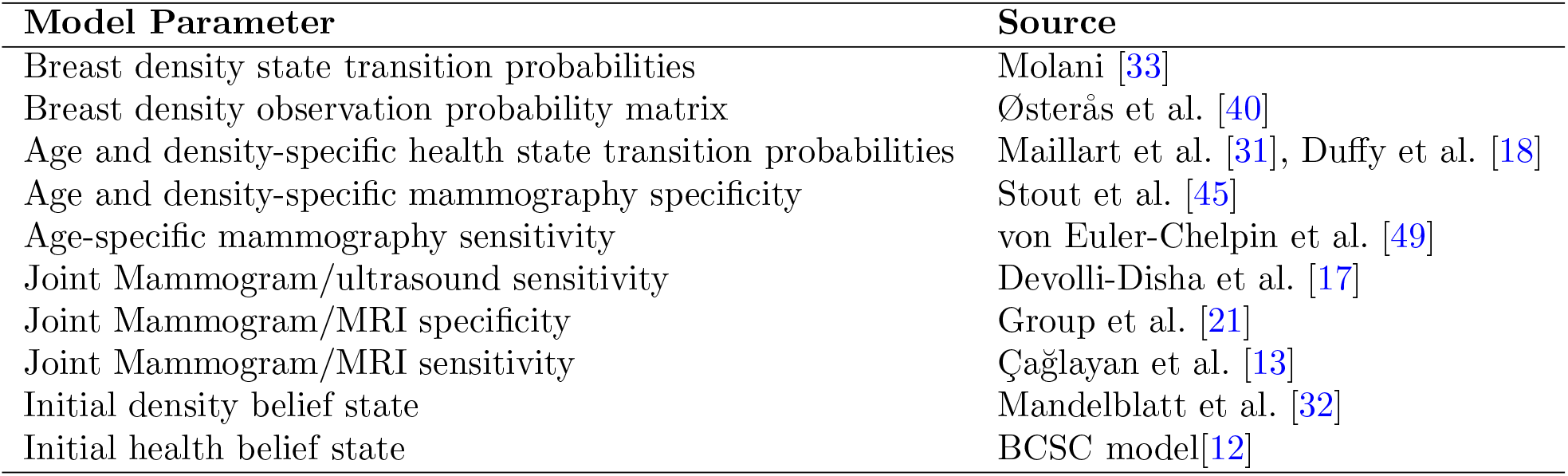
Source of model inputs and parameters estimation

| Model Parameter | Source |
| --- | --- |
| Breast density state transition probabilities | Molani [33] |
| Breast density observation probability matrix | Østerås et al. [40] |
| Age and density-specific health state transition probabilities | Maillart et al. [31], Duffy et al. [18] |
| Age and density-specific mammography specificity | Stout et al. [45] |
| Age-specific mammography sensitivity | von Euler-Chelpin et al. [49] |
| Joint Mammogram/ultrasound sensitivity | Devolli-Disha et al. [17] |
| Joint Mammogram/MRI specificity | Group et al. [21] |
| Joint Mammogram/MRI sensitivity | Çağlayan et al. [13] |
| Initial density belief state | Mandelblatt et al. [32] |
| Initial health belief state | BCSC model[12] |

### 3.1 Transition probabilities

As discussed earlier, previous studies have shown that mammographic breast density is associated with increased breast cancer risk. We estimate the age-specific and density-specific transition probabilities of the breast cancer natural history model by adjusting previously estimated transition probabilities by Maillart et al. [31] using different odds ratios comparing the risk of development and progression of breast cancer in different breast density classes. In our baseline analysis, we use odds ratio of 3.73, which is the midpoint of the odds ratio range (minimum and maximum OR values of 1.46 and 6), reported in the literature ([18], [22]). We will perform a sensitivity analysis to investigate the impact of a change in odds ratio on results in Section 4.1. To calculate the density-specific health transition probabilities, we adjust disease development and progression probabilities using the odds ratios of breast cancer risk comparing high and low breast density patients and the proportion of women in low and high breast density class in the U.S. For details, please see Appendix A.

We estimate breast density transition probabilities using longitudinal mammography screening data from Louisiana Cancer Prevention and Control Programs [28]. The dataset contains 436 patients with longitudinal mammogram screening data (including breast density assessments) between 2016 and 2020. Patients in the dataset are grouped into two different age categories of 40-54 and 55+ to capture the impact of age and menopausal status on breast density, as previous studies have shown a significant dependency between the menopausal status and breast tissue density [19]. Assuming that breast density is partially observable and missing observations are ignorable, we estimate the transition probabilities using the Baum-Welch method. Note that in the ignorable missingness mechanism, the probability of missingness depends only on the observed values and not the missing values [43].

### 3.2 Observations probabilities

We estimate the breast density information matrix for an average-skilled radiologist using a previous study by østerås et al. [40]. In their study, a number of radiologists interpreted 537 mammogram images and reported their density classifications. The radiologists’ classification results were then compared with the volumetric breast density obtained from a commercially available software (Quantra). They reported that in 87% of the cases the clinical interpretation agreed with radiologist reports. We also consider information matrices reflecting a perfect radiologist and radiologists who always downgrade (to minimize reporting requirements) and upgrade (to minimize liability) density classifications.

The health information matrices are estimated using screening tests sensitivity and specificity values. Specificity is defined as the probability of receiving a negative result when the patient is in cancer-free stage (i.e., true negative), and sensitivity is the probability of receiving a positive result when the patient is in a cancer state (i.e., true positive). Specifically, let *sens*_*t*_(*a*|*s* = (*h, d*)) denote the sensitivity of action *a* (*a ∈* {*M, B*}) when the patient is in cancer state *s* = (*h, d*), *h ∈* {1, 2} and *spec*_*t*_(*a*|*d*) denote the specificity of action *a* when the patient is in breast density class *d* at time *t*. The health information matrix elements for action *a* can be calculated as follows.

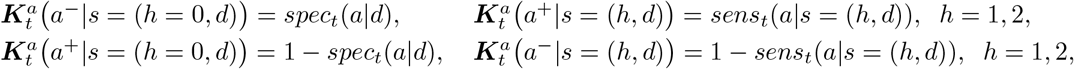

We use the cancer stage and density specific sensitivity and specificity of mammography provided in von Euler-Chelpin et al. [49] and Stout et al. [45], respectively. The sensitivity of joint mammogram/ultrasound and mammogram/MRI are adopted from Devolli-Disha et al. [17] and Ç ağlayan et al. [13], respectively. We use the specificity of joint mammogram/MRI from Group et al. [21].

### 3.3 Initial belief state

Initial health belief state is estimated using the Breast Cancer Surveillance Consortium (BCSC) risk model [12]. BCSC risk model estimates advanced breast cancer risk based on age, race, family history of breast cancer, history of a breast biopsy, and BI-RADS breast density [12]. To estimate the early breast cancer risk, we use the breast cancer stage distribution by race reported by the ACS [4], and the race distribution in the U.S. [46]. The ratio of early to advanced breast cancer cases among women in the U.S. is estimated as 1.78. The initial breast density distribution for the general population is adopted from Mandelblatt et al. [32]. We combine health and density initial belief (at age 45) to calculate the patient’s initial belief state. That is, the probability that a patient is in state *s* = (*h, d*) at the beginning of the screening horizon is

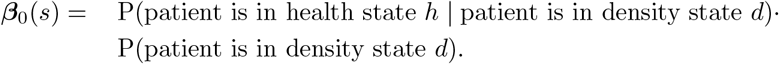

### 3.4 Model validation

To validate the estimated parameters, we calculate i) the lifetime risk of developing breast cancer from the proposed model, ii) five-year and ten-year risks of developing breast cancer from the proposed model, and iii) lifetime mortality risk of breast cancer with some adjustments to the model proposed by Molani et al. [34] to incorporate breast density.

Our estimation of the lifetime breast cancer risk (12.37% for the general population) is close to the reported ACS risk of 1 in 8 women (12.5%) [5]. More specifically, we estimate the lifetime risk of developing breast cancer as 7.56%, 17.18%, and 12.37% for women with low and high breast density at the beginning of the decision horizon, and for the general population (based on the average density), respectively.

The derived five-year and ten-year breast cancer risks are compared with the associated risks obtained from the BCSC risk assessment tool [12]. The estimated five-year risk of breast cancer using the BCSC risk assessment tool at age 60 and 70 are 1.68% and 2.00%, which are comparable with our estimations of 1.59% and 2.37%. Moreover, the ten-year breast cancer risks estimated using the BCSC risk tool are 2.51%, 3.45%, and 3.80% at age 50, 60, and 70. These are comparable with our corresponding estimated risks of 2.17%, 3.76%, and 4.03%, respectively.

We estimate the lifetime breast cancer mortality risk under several screening policies (see Table 2) for the general population (average-risk women). The average of the lifetime mortality risks across all the screening policies equals 2.88% that is comparable with the reported ACS risk of 1 in 38 women (2.63%) [5]. Note that we report the average lifetime breast cancer mortality risk across different policies (screening frequencies) to implicitly account for the variation in the breast cancer screening frequencies (adherence) for women in the U.S. Specifically, the estimated mortality risk for the two major screening policies in the U.S. are estimated as 1.06% for the ACS policy with switching screening intervals and 3.07% for the USPSTF policy.

**Table 2:** Screening policies considered in the numerical analyses

| Policy ID | Institution/Policy | Start age | End age | Screening Interval |  |  |  |
| --- | --- | --- | --- | --- | --- | --- | --- |
|  |  |  |  | 40-44 | 45-49 | 50-54 | 55+ |
| P1 | The annual option of the ACS policy (2015) | 45 | 75 | NA | 1 | 1 | 1 |
| P2 | The ACS policy with switching interval (2015) | 45 | 75 | NA | 1 | 1 | 2 |
| P3 | Biennial screening between age 45 and 75 | 45 | 75 | NA | 2 | 2 | 2 |
| P4 | Triennial screening between age 45 and 75 | 45 | 75 | NA | 3 | 3 | 3 |
| P5 | USPSTF (2016), AAFP (2016), France and Netherlands | 50 | 74 | NA | NA | 2 | 2 |
| P6 | Belgium, Denmark, Finland, Germany, Ireland, Poland and Spain | 50 | 69 | NA | NA | 2 | 2 |
| P7 | United Kingdom | 50 | 70 | NA | NA | 3 | 3 |

## 4 Numerical analyses

In this section, we evaluate the efficacy of supplemental screening and the impact of radiologists’ bias on patients outcomes for some of the in-practice screening policies. Table 2 presents the policies, denoted by P1 through P7, evaluated in this study. The screening policies recommended by the two major U.S. health agencies, the ACS and the USPSTF, along with some screening guidelines in the European countries are evaluated. These policies differ in terms of recommended screening starting and stopping ages and the screening interval. Biennial and triennial screening with the starting and stopping ages of 45 and 75 are also assessed. Additionally, we consider *do nothing* (DN) policy with no recommended screening test in a patient’s lifetime.

Per the breast density notification laws, in our numerical analyses, patients breast density are classified into two classes of low and high density where the former includes BI-RADs density classification of almost entirely fatty and scattered fibroglandular and the latter includes heterogeneously dense and extremely dense classes. Classifying patients into two density groups also reduces the computational complexity, especially for policies with a high number of prescribed screenings. Recall that we use sample-path enumeration approach for policy evaluation. This means that for policy P1, considering 4 possible density observations results in over 4^30^ sample paths which makes the policy evaluation computationally infeasible. If one wishes to consider four density classes, POMC simulation can be used for policy evaluation.

We consider 4 different radiologist types: radiologist with minimizing reporting requirement behavior (type 1), average-skilled radiologist (type 2), perfect radiologist (type 3), and radiologist with minimizing liability behavior (type 4). Radiologists type 1 and 4 always downgrade and upgrade patients’ breast density categories, respectively. Note that under the radiologist type 1, the patient never undergoes a supplemental test while under radiologist type 4, all screening mammograms are followed by a supplemental test. Radiologist type 3 (perfect radiologist) classifies breast density with 100% accuracy and radiologist type 2 (average-skilled) has 13% misclassification probability [40], as discussed in Section 3.2.

Four patient cases differing in breast cancer risk characteristics including race, breast density, breast cancer family history, and biopsy history are considered. The initial belief for these patients are calculated using the BCSC risk model [12]. These cases are as follows:

***Case 1***: A 45-year-old white woman with no breast cancer family history or prior biopsy. It is assumed that this case is in density class 1 at age 45. This patient is considered to be a low-risk case with initial estimated early and advanced breast cancer risks of 0.28% and 0.16%.

***Case 2***: All risk factors for this case are similar to *Case 1*, except for the initial breast density which is assumed to be extremely dense. This patient’s risks of being in early and advanced breast cancer states at age 45 are estimated as 1.19% and 0.67%, respectively.

***Case 3***: A 45-year-old white woman with a family history of breast cancer and prior biopsy and breast density class of 4. The patient’s estimated risks of early and advanced breast cancer at age 45 are estimated as 16.38% and 9.2%, respectively.

***Case 4***: This case represents the general (average-risk) population. The estimated risks of early and advanced breast cancer for the average-risk population at age 45 are 1.67% and 0.94%, respectively [36]. We estimate the initial belief state for this case using the breast density distribution of the women population in the U.S. provided by [32].

Figures 2 and 3 present the probability of detecting cancer in early and advanced states and the expected number of supplemental screening tests for different cases and under different radiologist types when ultrasound and MRI are used as supplemental screening tests, respectively. Note that under radiologists with minimizing reporting requirement behavior, patients only undergo mammogram tests. Obviously, more aggressive screening strategies are more likely to detect cancer in early states, where there is a higher chance of survival. That is, 1) the ACS policy with fixed screening intervals has the highest probability of detecting cancer in early states for all four cases and all radiologist types, and 2) under radiologist type 4 where patients always undergo supplemental screenings, patients receive the highest probability of being detected in early cancer states.

**Figure 2:**
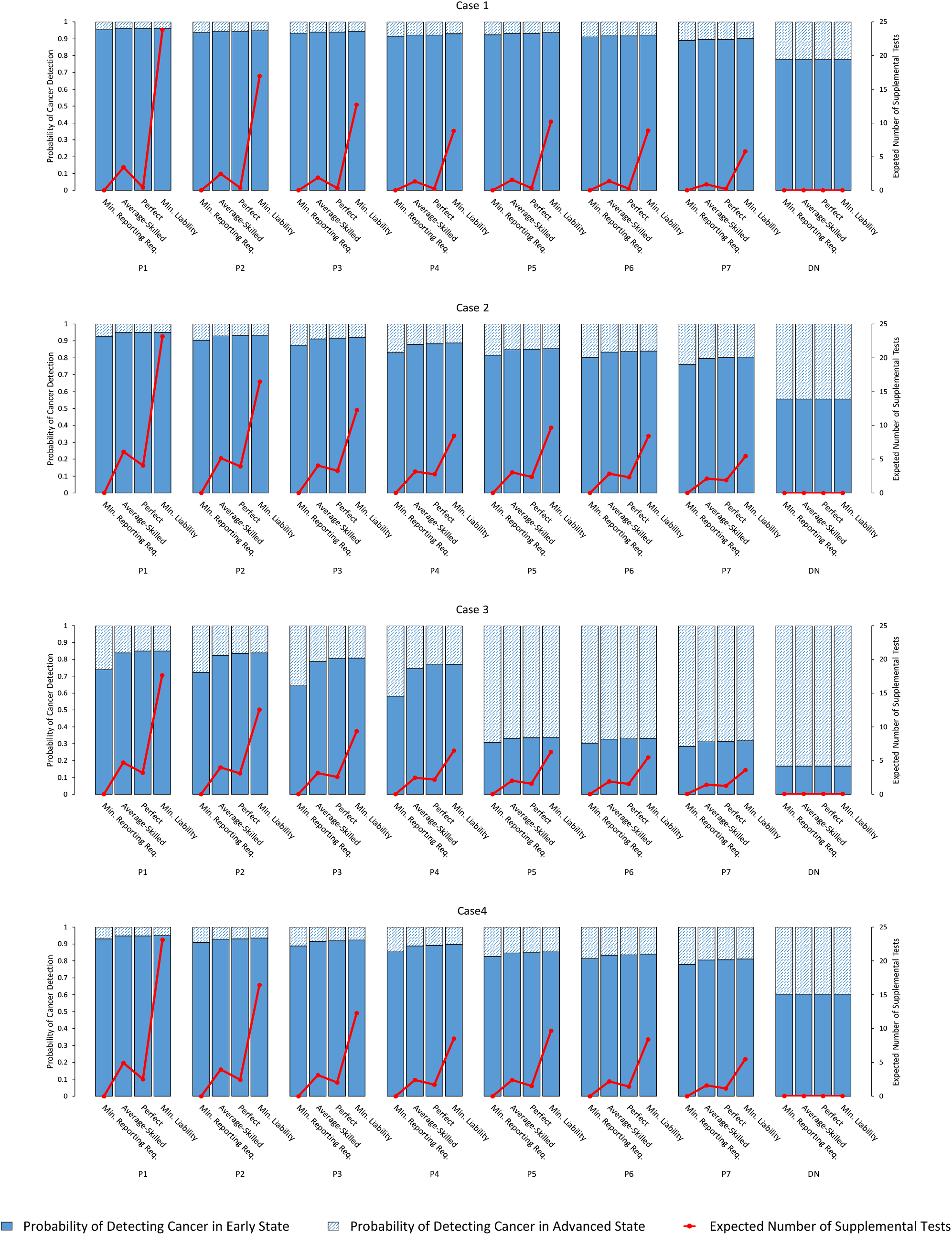
The conditional probabilities of detecting cancer in early and advanced states and the expected number of supplemental tests under different radiologist types–supplemental test: *ultrasound*.

**Figure 3:**
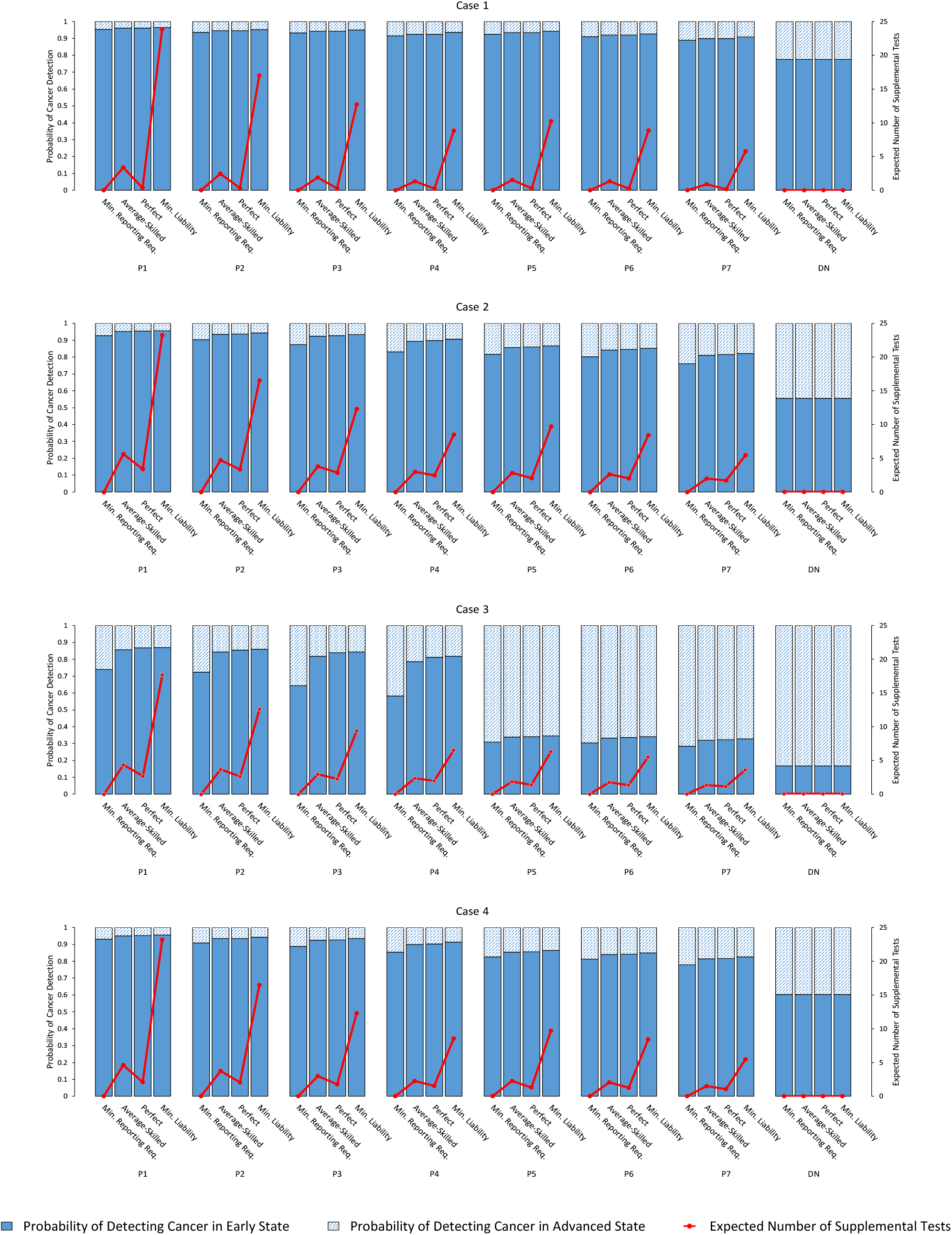
The conditional probabilities of detecting cancer in early and advanced states and the expected number of supplemental tests under different radiologist types–supplemental test: *MRI*.

Moreover, the results suggest that in terms of the probability of detecting cancer in early states, the difference in the performance of perfect and average-skilled radiologists is very small. In fact, the performances of perfect and average-skilled radiologists are very close to the performance of radiologist type 4. However, note that the radiologist type 4 impose a significantly higher number of supplemental screening tests on patients. For example, for *Case 1*, the expected number of supplemental tests that she undergoes are 23.85 and 16.97 under the two ACS policies. Note that this case does not really benefit from supplemental screening since she most likely remains in low breast density in her lifetime. Under the perfect radiologists, the corresponding expected number of supplemental screening tests are 0.41 and 0.36. Note that this implies that the patient undergoes 23.44 and 16.61 unnecessary supplemental tests under radiologist type 4 which adversely affects her quality of life. Under the average-skilled radiologist, the expected number of supplemental tests are 3.41 and 2.48 for the two ACS policies, which suggests that the unnecessary number of supplemental tests are 3.00 and 2.12, respectively. The expected number of unnecessary supplemental tests are smaller under the other screening policies as they are less aggressive. This implies that average-skilled radiologists’ performance is very close to perfect radiologists’ performance when comparing the probability of detecting cancer in the early states. However, in terms of the expected number of supplemental tests, the difference might be significant (depending on the aggressiveness of screening policies).

The differences in the performance of radiologists become more evident as the patient’s risk increases. That is, for *Case 1* and *Case 3*, the differences are at their lowest and highest level, respectively. Specifically, for *Case 1* and under *ultrasound* as the supplemental test, the probability of detecting cancer in early states increases by only 0.63%, and 1.11% for the two ACS policies (P1 and P2) when going from radiologist type 1 to type 4. For *Case 3*, however, the corresponding increases are 11.04% and 11.52%, respectively. *Case 2* and *4* fall in between *Case 1* and *3*.

This also implies that the efficacy of supplemental screening highly depends on the patients’ overall breast cancer risk and not only their breast density. For patients with a lower risk (e.g., *Case 1*), the benefit of undergoing supplemental screening is minimal, as discussed above. For *Case 2* with all risk factors similar to *Case 1* but breast density, we observe an increase in the probability of cancer detection in early states when the patient undergoes supplemental tests. For instance, under mammogram only policy (radiologist type 1), the early detection probability is 92.75% for policy P1 and this probability increases to 94.89% and 95.00% under radiologist types 2 and 3 who recommend the patient undergo ultrasound test as needed, per breast density notification laws. For *Case 3*, however, undergoing supplemental screening provides a significantly higher benefit, as discussed above.

Comparing MRI and ultrasound, we observe that MRI always results in a higher probability of detection in early states as it is more sensitive than ultrasound. The difference, although, is negligible, especially for low-risk cases. The highest difference in the performance of MRI and ultrasound occurs for *Case 3*. For this case, under the average-killed radiologist and biennial and triennial screening policies, using MRI results in the corresponding early detection probabilities of 81.85% and 78.56%, as compared with 78.76% and 74.61% when undergoing ultrasound tests. Additionally, MRI results in a slightly lower expected number of the supplemental tests compared to ultrasound since it has slightly lower false negatives (higher sensitivity). Note that MRI is a more aggressive test and therefore it might be beneficial to be used only for the cases with higher risk such as *Case 3*.

Generally, the results suggest that the policies with the starting age of 45 outperform those with the starting age of 50. That is because breast cancer is more aggressive at younger ages. The impact of starting age, in particular, and screening policy, in general, on detecting cancer in early states is specifically very evident for *Case 3*. This implies that the probability of detecting cancer in early states is more impacted by the policy type and patient risk than the radiologist’s type.

In summary, the results show that 1) breast density is not a sufficient factor when administering supplemental screening as the increases in probabilities of detecting cancer in early states in all cases, except for *Case 3*, are very negligible among different radiologist types for any given policy. Other risk factors must be taken into account when recommending a supplemental screening (as shown in *Case 3*). 2) In terms of the probability of detecting cancer in early states, average and perfect radiologists’ performance are very similar and comparable to the performance of radiologist type 4. Additionally, if the patient is not a high-risk case, radiologist type 1 performance is also comparable to the other radiologist types. This implies that radiologist type impact is not as significant as other factors such as the patient risk factors and screening frequency.

### 4.1 Sensitivity analyses

In this section, we conduct sensitivity analyses on the 1) odds ratio (OR) of breast cancer risk comparing women in different density classes and 2) sensitivity and specificity of supplemental screening test and the probability of cancer showing symptoms. These parameters are selected due to the variability in their reported values in the literature. We consider 4 different ORs, excluding the baseline OR. In the second part of sensitivity analyses, we consider 18 different combinations for the sensitivity and specificity of the supplemental test and the probability of cancer showing symptoms. In total, for each patient case, we evaluate the outcomes under 832 settings.

#### 4.1.1 Odds ratios

Based on the previous studies, the odds ratio for developing breast cancer for the high breast density patients compared with low breast density patients ranges from 1.46 to 6.0 [18, 22]. We use the midpoint value (OR=3.73) in our baseline analyses, presented in Figures 2 and 3. Here, we consider OR values of 1.46, 2.595, 4.865, and 6. These ORs are selected to include minimum and maximum values reported in the literature, as well as the midpoint values of the intervals formed by these values and the baseline OR. Clearly, as the OR increases, the breast cancer risk gap between women with high and low breast density increases.

Figure 4 presents the change in the probability of detecting cancer in early states caused by a change in OR values, when compared to the baseline. Note that negative and positive changes present a decreased and an increased probability of detection in early cancer states, respectively.

**Figure 4:**
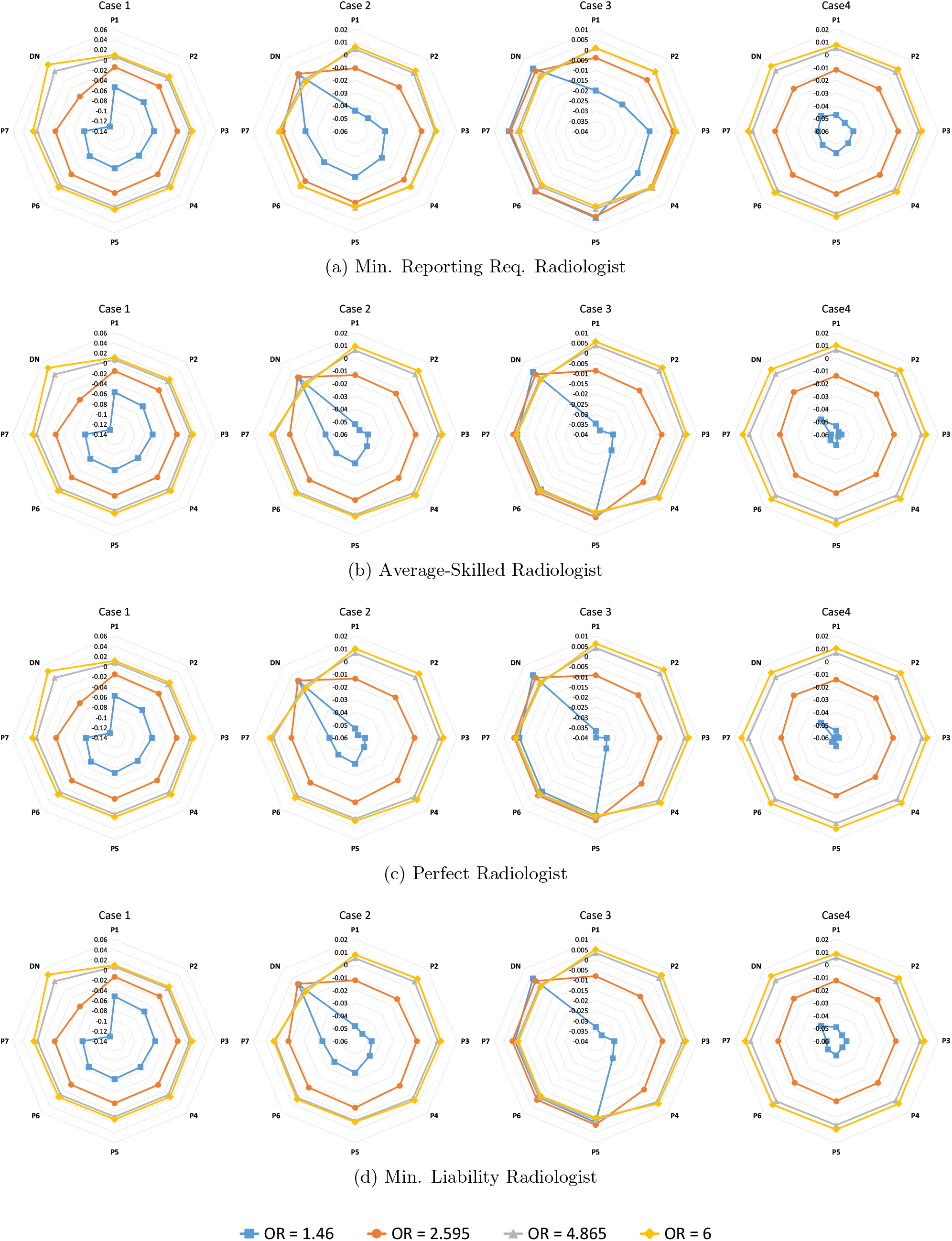
Results of sensitivity analyses on the odds ratio of breast cancer comparing low and high breast density classes. Note that negative/positive values imply a decreased/increased probability of detecting cancer in early states compared with the baseline (*OR* = 3.73).

Under the maximum OR value (*OR* = 6), we observe an average absolute change of 1.08% across all cases, radiologist types and policies, with a maximum of 4.5% (under *Case 1*, radiologist type 1, and *do nothing* policy). Under increased OR, patients with low breast density (e.g., *Case 1* at age 45) carry lower breast cancer incidence and progression rates compared to the baseline. On the other hand, patients with high breast density (e.g., *Case 2* at age 45) carry higher breast cancer risk. Interestingly, the results suggest that for both of these cases, the probability of early detection increases (except for *Case 2* under *do nothing* policy.) This is expected for *Case 1* as this case starts and most likely remains in density class 1 in her lifetime. For *Case 2*, the increase in detection probability seems counterintuitive since this case has an increased breast cancer risk due to occupying density class 4. However, note that the increase in early detection probability is very negligible and due to the probable transition of this patient to a low-density class over the course of a few epochs. Note that, based on our data and previous studies, breast density stochastically decreases over time [27, 48, 51]. For the other two cases, the changes are very negligible, especially for *Case 3*. Obviously, under the OR value of 4.865, the changes are smaller but follow similar patterns.

With a decreased OR, we observe higher changes in the probability of early detection. Specifically, under OR value of 1.46, we observe a maximum change of 12.72% (for *Case 1* under *do nothing* policy). In general, we observe a decrease in probability of early detection for *Case 1, 2*, and *4* (except for *do nothing* policy for *Case 2*). Note that with a decreased OR, patients with lower breast density carry a higher risk compared with the baseline which results in a decrease in the probability of early detection. Note that for *Case 2*, although she starts in high-density class, it is very likely that she will transition to low density in a few epochs. Under *do nothing* policy for *Case 2*, we observe a very negligible change in early detection probability. Note that the small change in this case is mainly contributed by the patient’s belief of being in cancer states. The patient’s cancer belief under this policy is generally higher due to the lack of screening tests and consequent less informative risk adjustments of the patient. This is also true for *Case 3* whose change in early detection probability is very negligible for *do nothing* policy and screening policies starting at age 50. Note that for policies with a starting screening age of 50, this patient whose risk is already high at age 45 (due to her risk factors) does not get screening opportunities to detected cancer in early states before cancer progresses to advanced states.

Generally, with increased/decreased OR, we observe an increase/decrease in early detection probability. The magnitudes of changes, however, vary across different patients and screening policies. The results, in general, are consistent and prompt comparable conclusions with those derived under the baseline OR.

#### 4.1.2 Supplemental test accuracy and probability of cancer symptoms

We consider 3 different levels of sensitivity and 3 different levels of specificity for the supplemental screening test. We also consider 2 different levels for the probability of cancer showing symptoms. Using a full factorial design, we have 18 different combinations for these parameters. Specifically, we consider 1) joint sensitivity of mammogram and ultrasound decreased by 5%, 2) joint sensitivity of mammogram and MRI increased by 5%, and 3) the midpoint of the interval formed by the joint mammogram/ultrasound and mammogram/MRI sensitivity values in the baseline. Similarly, we calculate three levels for supplemental screening specificity. We consider 5% increase and decrease in the baseline probability of cancer showing symptoms.

Figure 5 presents the change in the probability of detecting cancer in early states for *Case 3* under an average-skilled radiologist. We present the results only for *Case 3* for brevity; however, note that in general, sensitivities to parameter changes are smaller for the other cases. For example, the maximum change in the probability of detecting cancer in early states for *Case 1* is only 1.57% when compared with the baseline, while the maximum change for *Case 3* is 8.44%.

**Figure 5:**
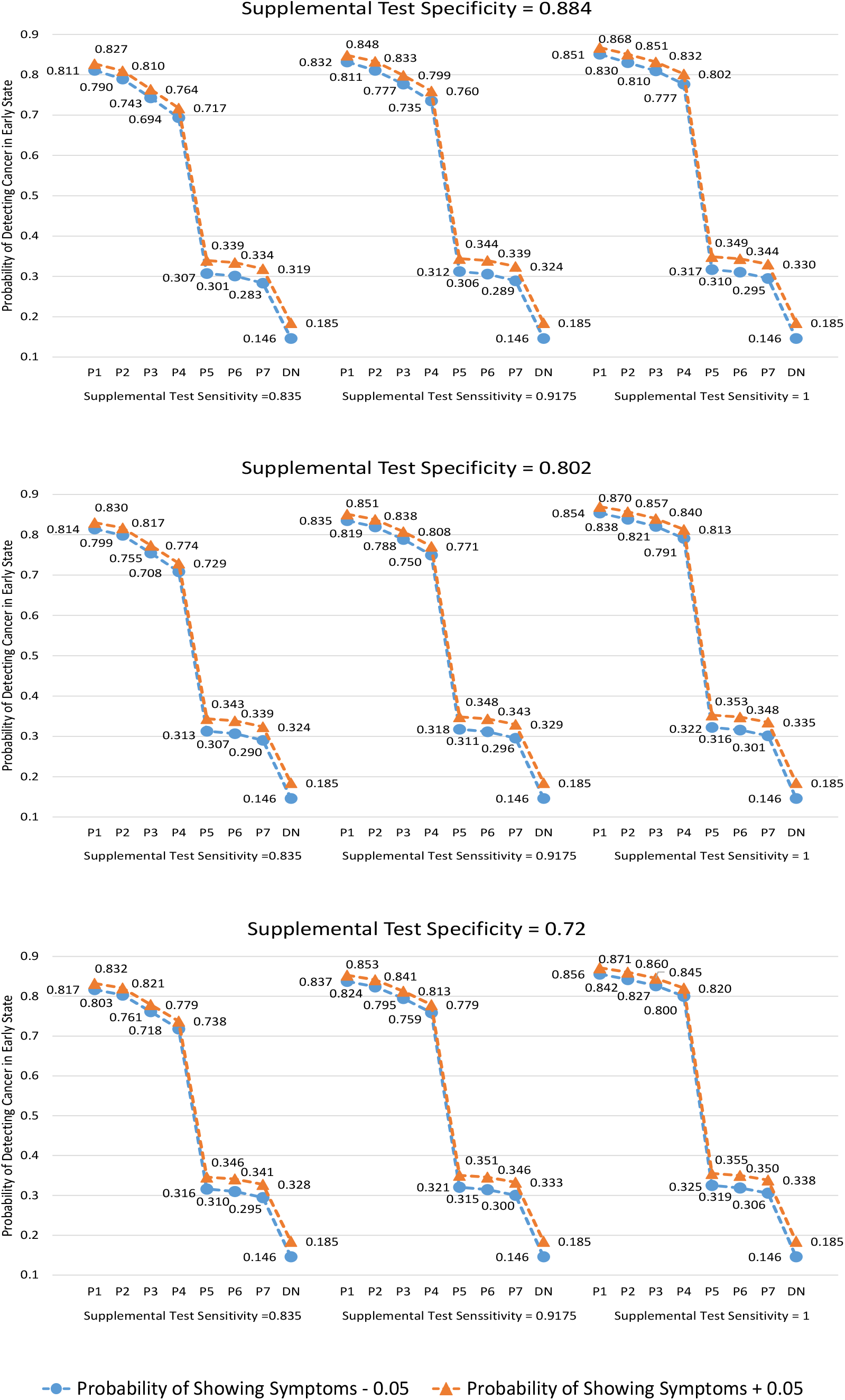
Results of sensitivity analyses on the joint mammogram and the supplemental test sensitivity and specificity and the probability of cancer showing symptoms: *Case 3* and average-skilled radiologist.

Obviously, the change in sensitivity of testing has the highest impact on the probabilities of early detection. As the test sensitivity increases, the probability of detecting cancer in early states increases. The increase in early detection probability is smaller for policies with more frequent screening policies as more frequent screenings compensate for possible false negatives. The increase is at the highest for policy P4, due to the longer intervals between subsequent recommended screening tests (3 years). Note that for policy P7, although following the same screening frequency, the change is very small due to the delayed screening starting age. That is, it is very likely that cancer remains undetected and grows to an advanced state between ages 45 and 50 which consequently decreases the chance of detecting cancer in early states. Recall that *Case 3* is a high-risk case and has aggressive breast cancer.

Similarly, increasing the probability of showing symptoms results in an increase in the probability of detecting cancer in early states. The highest impact is associated with less aggressive policies. For instance, for *Case 3* and under *do nothing* policy, we observe an increase of 3.86% in the probability of early detection when increasing the probability of showing symptoms. On the other hand, for the ACS policy with fixed screening intervals (P1), we observe the lowest sensitivity to the probability of showing symptoms, with an average increase of 1.43% across different sensitivity analyses combinations considered here.

The specificity of screening tests has an opposite impact on the probability of detecting cancer in early states. That is, increased joint specificity causes a decrease in the probability of early detection. This happens as with increased joint specificity, the number of false-positive observations, resulting in consequent biopsies, decreases. Recall that biopsies are perfect and determine with certainty that the patient is cancer-free. This causes an overall decrease in the belief that the patient is in cancer states and the probability of detecting cancer in the early states.

In summary, the sensitivity of joint mammogram and the supplemental test has the largest and the specificity of joint mammogram and the supplemental test has the smallest impact on the probability of early detection. The conclusions in the baseline analyses still hold. That is, as long as a supplemental test is administered for a patient (radiologist types 2–4), a bias in radiologist’s classification has a negligible impact on the probability of detecting cancer in early states. Moreover, breast density should not be the sole determining factor as to whether a patient should be referred to supplemental screening tests. Other breast cancer risk factors and frequency of screening tests should be considered when making such referrals.

## 5 Conclusion

Breast density is associated with increased breast cancer risk and decreased mammography screening sensitivity. To promote early breast cancer detection in women with high breast density, breast density notification laws have been enacted in 38 states. The laws, however, have caused controversial debates on 1) whether supplemental screening improves patients’ outcomes and 2) the impact of radiologists’ bias in breast density classification on patient outcomes.

In this study, we develop a POMC model, incorporating both patients’ health and breast density dynamics, to investigate the impact of supplemental screening tests and the role of radiologists’ bias on a patients’ health outcomes. Specifically, we consider the conditional probability of detecting breast cancer in early and advanced states given the patient develops breast cancer in her lifetime. We consider the expected number of supplemental tests a patient undergoes in her lifetime as another patients’ outcome.

Our results indicate that 1) breast density should not be the only risk factor when referring a patient to supplemental screening and 2) the radiologist’s bias may affect the efficacy of supplemental screening. Specifically, patients’ outcomes may be significantly affected under radiologists who consistently upgrade or downgrade patients’ breast density. However, the bias introduced by an average-skilled radiologist may not significantly affect the patients’ outcomes. There are some packages, such as EIZO, CADx [26], that can assist radiologists with breast density classification. These packages leverage artificial intelligence and machine learning tools to classify mammogram images into different breast density classes.

Given that screening technologies are continuously advancing, a future research direction is to analyze the impact of emerging technologies (e.g., tomosynthesis) on the necessity of supplemental screening tests. Moreover, patients’ adherence is a very influential factor in the effectiveness of screening policy and patients’ outcomes. A possible future work is to incorporate this factor.

## Data Availability

No data available to report.

## Appendix A Estimation of transition probabilities

Let 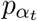 be the proportion of women in low breast density at age *α*_*t*_ associated with time period *t*. Let *I*_*t*_ denote the general breast cancer incidence probability (i.e., probability of going from cancer-free state to early breast cancer state) at time *t*. Moreover, let 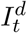 represent the incidence probability for patients in breast density class *d* at time *t*. We calculate the incidence probability for low and high breast density at time *t*, denoted by 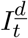 and 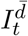 respectively, using the following set of equations.

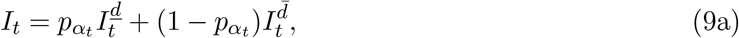

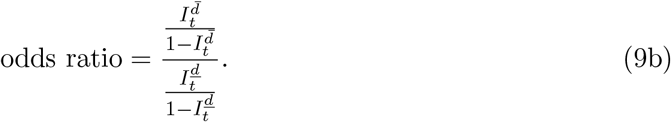

Therefore, the core transition probability ***P***_***t***_ (*s* ^*′*^ = (1, *d* ^*′*^)|*s* = (0, *d*)) is calculated as *Pr*(transition from density state *d* to *d* ^*′*^) 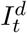, where breast density transition probabilities are adopted from Molani [33]. The cancer progression probabilities is calculated using a similar approach.

